# Lower Physical Activity Modifies the Association between Perceived Fatigability and Executive Function but not Memory: The Study of Muscle, Mobility and Aging (SOMMA)

**DOI:** 10.1101/2023.11.06.23298173

**Authors:** Benjamin T. Schumacher, Caterina Rosano, Yujia (Susanna) Qiao, Andrea L. Rosso, Peggy M. Cawthon, Kyle D. Moored, Steven R. Cummings, Stephen B. Kritchevsky, Nancy W. Glynn

**Affiliations:** Department of Epidemiology, School of Public Health, University of Pittsburgh, PA, USA; Department of Epidemiology and Biostatistics, University of California, San Francisco, CA, USA; Research Institute, California Pacific Medical Center, San Francisco, CA, USA; Department of Mental Health, Johns Hopkins Bloomberg School of Public Health, Baltimore, MD, USA; Wake Forest School of Medicine, Gerontology and Geriatric Medicine, Winston-Salem, NC, USA

**Keywords:** fatigue, mental fatigability, physical fatigability, cognition, cognitive function

## Abstract

**OBJECTIVE:** Emerging evidence shows that perceived fatigability—the quantification of vulnerability to fatigue in relation to specific intensity and duration of activities—may be associated with cognitive function. We sought to quantify associations with multiple domains of cognitive function and the role of physical activity (PA).

**METHODS:** SOMMA participants completed the Pittsburgh Fatigability Scale (PFS) Physical and Mental subscales (each range 0–50; higher scores=greater fatigability) and three cognitive function assessments [Digit Symbol Substitution Test (DSST), executive function; Montreal Cognitive Assessment (MoCA), general function; and California Verbal Learning Test (CVLT), memory]. Linear regression quantified associations cross-sectionally between each PFS subscale and cognitive assessment scores adjusting for covariates. Effect modification by volume and intensity of accelerometer-measured PA was assessed.

**RESULTS:** In 873 participants (59.2% women; age 76.3±5.0; 85% White), mean PFS Physical, Mental, and DSST scores were 15.8±8.7, 7.7±7.8, and 55.4±13.7. After adjustments, for each 4-point higher PFS Physical and 3-point higher PFS Mental, participants had nearly one fewer correct DSST items [β coefficient and 95% confidence interval for PFS Physical: -0.69 (-1.09, - 0.29); PFS Mental: -0.64 (-0.97, -0.30)]. Volume and intensity of PA modified the association of PFS Mental and DSST (*P_interactions_*<0.01). All associations were strongest in those with the lowest volume and intensity of PA. PFS was not associated with MoCA or CVLT.

**DISCUSSION:** Greater perceived fatigability may be associated with poorer executive function, but not memory. Individuals with greater perceived fatigability, particularly those less active, might benefit from interventions that reduce fatigability and may beneficially influence cognitive function.

## INTRODUCTION

Cognitive function is the quantification of performance across multiple cognitive domains, including, but not limited to, memory, language, perception, attention, executive function, learning, understanding, awareness, reasoning, and judgment (Fisher et al., 2019; Lezak et al., 2004). As these domains are associated with activities of daily living, preserving cognitive function is a key component for older adults to maintain independence (“2022 Alzheimer’s Disease Facts and Figures,” 2022). As the population of Americans over age 60 has been growing and is expected to rapidly increase over the next 30 years (U.S. Department of Health and Human Services, Administration for Community Living, 2021), understanding the associations between cognitive function and potentially modifiable up-stream health behaviors is a critical need to ensure older adults maintain prolonged independence, better cognitive function, and higher quality of life.

Perceived fatigability is a characteristic that measures an individual’s vulnerability to fatigue relative to activities of specific intensity and duration. Perceived physical fatigability captures capacity and effort, while perceived mental fatigability assesses mental tiredness (e.g., psychological, emotional, and cognitive). Perceived fatigability has been established as a prognostic indicator of several age-related health outcomes (Glynn et al., 2022; Qiao et al., 2019; Simonsick et al., 2018). Further, there exists a growing body of evidence that greater perceived physical and mental fatigability are associated with lower brain grey matter volumes (Wasson et al., 2019) and poorer performance on the Digit Symbol Substitution Test (LaSorda et al., 2020; Renner et al., 2021), a measure of executive function and processing speed. As one’s cognitive function is crucial to maintaining independence, understanding the associations of perceived physical and mental fatigability with multiple cognitive function domains may inform the design of intervention programs aimed to reduce fatigability, maintain cognitive function, and contribute to the healthy longevity of older adults.

Previous work suggests that reductions in physical activity (PA) may precede and predict perceived fatigability (Qiao et al., 2021, 2022). Furthermore, strong evidence exists that higher amounts of PA can preserve/maintain better cognitive function (2018 Physical Activity Guidelines Advisory Committee, 2018; “2022 Alzheimer’s Disease Facts and Figures,” 2022). Therefore, it is important to examine whether PA confounds and/or modifies the association of perceived fatigability with cognitive function, as older adults may restrict the quantity or intensity of PA to avoid feelings of fatigue (Avlund et al., 1996; Eldadah, 2010).

We sought to address these research gaps by quantifying the associations of perceived physical and mental fatigability, as measured by the Pittsburgh Fatigability Scale (PFS) Physical and Mental subscales, with the Digit Symbol Substitution Test (DSST), Montreal Cognitive Assessment (MoCA), and/or the California Verbal Learning Test (CVLT) in 873 community-dwelling older adults. Due to the relation between PA and physical fatigability, and the well-established relation between PA and cognitive function, we hypothesized that perceived physical fatigability would be more strongly associated with each of the cognitive function assessments than perceived mental fatigability. Next, we evaluated how adjustment for accelerometer-measured PA would influence our models. We hypothesized that the adjustment for PA would attenuate the observed associations. Finally, in attempt to gain a deeper understanding of the role PA serves in the fatigability and cognitive function association, we assessed whether the associations were modified by PA. We hypothesized that those with higher levels of accelerometer-measured PA would have a weaker association relative to those with lower levels of PA.

## METHODS

### Study Participants

The Study of Muscle, Mobility and Aging (SOMMA) (https://sommaonline.ucsf.edu) is an on-going, prospective cohort study of 879 adults ≥ 70 years that seeks to understand the biologic mechanisms that underlie mobility decline. Details about eligibility, enrollment, and data collection in SOMMA have been published elsewhere (Cummings et al., 2023). At baseline, SOMMA participants must have been able to complete the 400-meter walk; those who appeared as they might not be able to complete the 400-meter walk at the in-person screening visit completed a short distance walk (4 meters) to ensure their walking speed was ≥ 0.6m/s. Additionally, the must have been free of life-threatening disease and have been able to undergo magnetic resonance imaging and tissue collection (Cummings et al., 2023).

All SOMMA participants provided written informed consent and the study was approved by WIRB-Copernicus Group (WCG) Institutional Review Board (study number 20180764).

### Measures

#### Exposures: Perceived Physical and Mental Fatigability from the Pittsburgh Fatigability Scale

The Pittsburgh Fatigability Scale (PFS) (Glynn et al., 2015) is a well-validated, questionnaire designed to separately measure perceived physical and mental fatigability (LaSorda et al., 2020; Renner et al., 2021). SOMMA participants rated on a scale from 0 (no fatigue) to 5 (extreme fatigue) the level of fatigue they expected they would experience after completing 10 different activities of various duration and intensities. The sum of the 10 items yields the PFS Physical and PFS Mental scores, both ranging from 0 to 50, with higher scores indicating greater perceived fatigability. PFS scores were imputed if 1-3 items were missing (n = 11 PFS Physical and n = 14 PFS Mental) (Cooper et al., 2019).

For descriptive and exploratory purposes, we assessed PFS scores in their continuous form and by severity strata: 0 – 4, 5 – 9, 10 – 14, 15 – 19, 20 – 24, and ≥25 for PFS Physical and 0 – 3, 4 – 7, 8 – 12, 13 – 15, 16 – 19, and ≥20 for PFS Mental, respectively (Cohen et al., 2021; Simonsick et al., 2018). Additionally, the prevalence of those with more severe perceived fatigability was calculated for PFS Physical (≥ 15) and PFS Mental (≥ 13) (Simonsick et al., 2018). The regression analyses in the present study used the clinically relevant 4-point and 3-point higher scores for PFS Physical and PFS Mental, respectively (Simonsick et al., 2018).

#### Outcomes: Cognitive Function Assessments

##### WAIS-III Digit Symbol Coding Subtest

The WAIS-III Digit Symbol Coding Subtest (licensed by NCS Pearson, Inc.; hereafter referred to as the Digit Symbol Substitution Test (DSST)) is a paper-and-pencil assessment where a participant is expected to substitute numbers with symbols according to a key (Jaeger, 2018). Scores on the DSST can range from 0 – 133 and represent the number of correct symbols substituted within two minutes, with a higher score indicating better cognitive function. The DSST assesses executive functioning, processing speed, motor speed, visuoperceptual functions, attention, and manual dexterity (Jaeger, 2018).

##### Montreal Cognitive Assessment (MoCA)

The MoCA is a dementia screening instrument that assesses the following cognitive domains: attention, concentration, executive function, memory, language, visuoconstructional skills, conceptual thinking, calculations, and orientation (Julayanont & Nasreddine, 2017; Nasreddine et al., 2005). SOMMA administered the MoCA in-person at the baseline visit. The MoCA has exhibited a high sensitivity and specificity for detecting cognitive impairment. MoCA scores can range from 0 – 30, with a higher score indicating better cognitive function.

##### California Verbal Learning Test-Second Edition, Short Form (CVLT-II SF)

Discussion on the extensive details on the methodology of the CVLT-II SF (licensed by NCS Pearson, Inc.; hereafter referred to as the CVLT) have been published elsewhere (Elwood, 1995). In brief, the CVLT consists of the participant being read a list of nine nouns from three categories (fruits, tools, clothing). After each set of nine nouns, the participant is asked to recall as many words as possible, in any order, to assess their immediate recall. During this immediate recall, any intrusion (a recalled word that was not in the original nine) and duplications (recalling a word more than once) are noted. This process is repeated four times. After 10 minutes, the participant is asked to recall as many words as possible, in any order, to assess their long-delay recall. Intrusions and duplications are noted. Finally, the participants are read a long list of words and asked whether each word was among the nine they were originally asked to memorize. For the purposes of these analyses, the number of items correctly recalled in the immediate recall task will be considered the CVLT Immediate Recall score (range 0 – 36) and the number of items correctly recalled in the long-delay recall task will be considered the CVLT Delayed Recall score (range 0 – 9). The CVLT assesses one’s verbal learning ability, semantic organization, intrusion, and clustering (Elwood, 1995).

### Covariates

Covariates included study site (University of Pittsburgh, Wake Forest School of Medicine), self-reported age, sex, race (White, non-White), educational attainment (high school or less, some college, college graduate, graduate school, or other), and marital status (married, no longer married, never married). We also included a modified version of the Rochester Epidemiology Project Multimorbidity Index (Goodman et al., 2013) that contained 11 age-related conditions, with one point added to the score for each condition the participant self-reported as part of their medical history (i.e. higher scores = more morbidity) (Espeland et al., 2017). The conditions considered were a self-report of a physician diagnosis of cancer (excluding nonmelanoma skin cancer), cardiac arrhythmia, chronic kidney disease, chronic obstructive pulmonary disease, coronary heart disease, congestive heart failure, dementia (zero for all participants), Center for Epidemiologic Studies Depression Scale (CES-D) (Radloff, 1977), diabetes, stroke, and aortic stenosis. Body mass index from measured height and weight was conceptualized as a potential mediator of the perceived fatigability and cognitive function association and was not included in these analyses except for descriptive purposes.

Additionally, the ActiGraph GT9X (ActiGraph, Pensacola, FL) was worn on the participant’s non-dominant wrist for 7 full days and collected raw accelerometer data at 80 hertz. We used two variables from the accelerometry data: 1) the average 24-hour total activity count (where an adherent wear day was defined as ≥ 17 hours of device wear time), without the low frequency extension filter, with imputation (Schrack et al., 2014) (i.e. total volume of PA), and 2) mean daily minutes of moderate-to-vigorous PA (MVPA), with imputation (i.e. intensity of PA).

### Statistical Analysis

Six participants were excluded for having missing perceived fatigability scores that could not be imputed for a sample size of N = 873. The final sample size varied slightly by cognitive assessment due to variation in the amount of missing data (n missing: MoCA = 11, DSST = 7, CVLT Immediate Recall = 16, and CVLT Delayed Recall = 23).

First, we assessed the baseline characteristics and cognitive function assessment scores across the PFS Physical severity strata using chi-square tests for categorical variables and analysis of variance (ANOVA) tests for continuous variables.

Next, multiple linear regression models were generated to estimate the associations between PFS Physical score (beta coefficient expressed per 4-point increment) and PFS Mental score (beta coefficient expressed per 3-point increment) and the three cognitive function assessments. In both approaches, Model 1 adjusted for age, sex, race, education, marital status, and site. Model 2 further adjusted for the multimorbidity index. To assess whether PA is a confounder of the perceived fatigability – cognitive function association, Models 3 and 4 introduced adjustments for total activity counts and mean daily minutes of MVPA, respectively.

Finally, we sought to assess whether the association of perceived fatigability and cognitive function association was modified by volume and intensity of physical activity. The average 24-hour total activity count and mean daily minutes of MVPA were included in product terms (PA*PFS) in unadjusted models. Effect modification was *a priori* considered significant at the *P* < 0.10 level. Models were stratified by tertile of the PA variable and fit following the same sequential adjustment strategy.

All associations, except for the interaction terms, were considered statistically significant if *P* < 0.05. All analyses were conducted in R version 4.1.0 (R Foundation for Statistical Computing, Vienna, Austria).

## RESULTS

### Sample Characteristics

The 873 participants were 59% women, 85% White, 13.3% Black, 0.7% Asian, 51% married, and 62% college educated or higher, and had mean (± standard deviation) age of 76.3 ± 5.0 years and BMI of 27.6 ± 4.6 kg/m^2^ (Table 1). The prevalence of more severe perceived physical (PFS Physical ≥ 15) and mental (PFS Mental ≥ 13) fatigability was 54.1% and 23.3%, respectively. Mean score for the DSST was 55.4 ± 13.7 (observed range: 7 – 95), MoCA was 24.8 ± 2.9 (observed range: 13 – 30), CVLT Immediate Recall was 25.5 ± 4.4 (observed range: 5 – 35), and CVLT Long Recall was 6.0 ± 2.0 (observed range: 0 – 9) (Table 1).

**Table 1.**
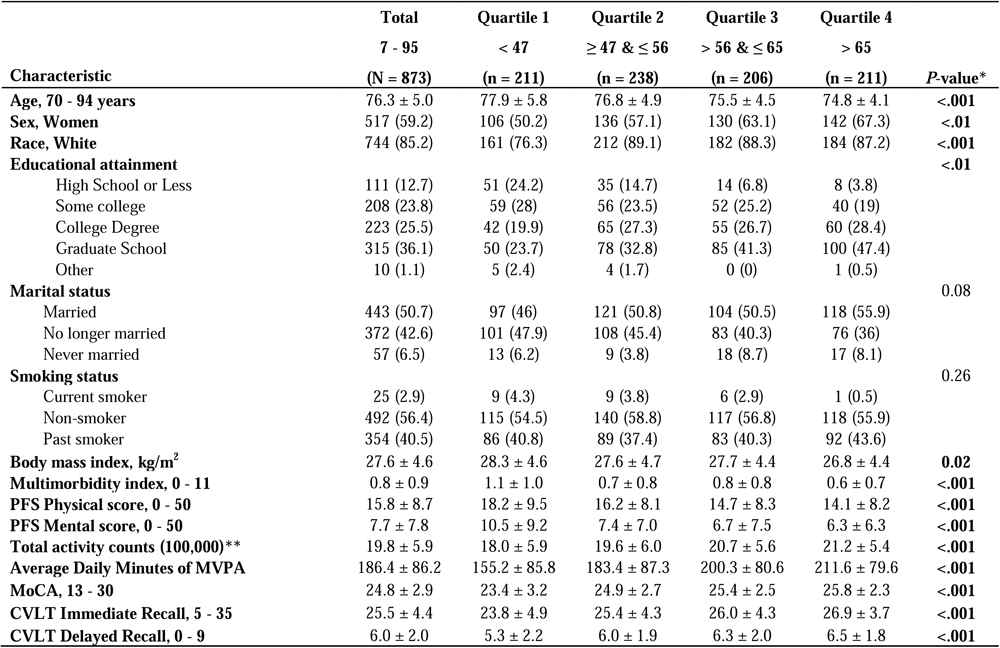
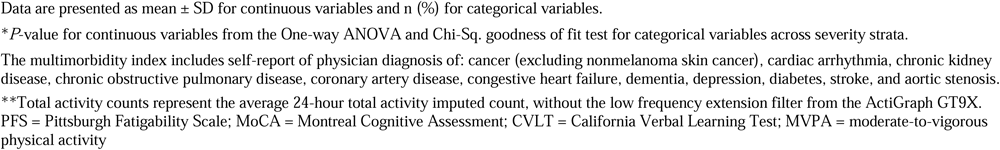
B aseline Participant Characteristics Stratified by Quartiles of the Digit Symbol Substitution Test: The Study of Muscle, Mobility and Aging (SOMMA) (N = 873)

|  | Total | Quartile 1 | Quartile 2 | Quartile 3 | Quartile 4 |  |
| --- | --- | --- | --- | --- | --- | --- |
|  | 7 - 95 | < 47 | ≥ 47 & ≤ 56 | > 56 & ≤ 65 | > 65 |  |
| Characteristic | (N = 873) | (n = 211) | (n = 238) | (n = 206) | (n = 211) | P-value* |
| **Age, 70 - 94 years** | 76.3 ± 5.0 | 77.9 ± 5.8 | 76.8 ± 4.9 | 75.5 ± 4.5 | 74.8 ± 4.1 | <.001 |
| **Sex, Women** | 517 (59.2) | 106 (50.2) | 136 (57.1) | 130 (63.1) | 142 (67.3) | <.01 |
| **Race, White** | 744 (85.2) | 161 (76.3) | 212 (89.1) | 182 (88.3) | 184 (87.2) | <.001 |
| **Educational attainment** |  |  |  |  |  | <.01 |
| High School or Less | 111 (12.7) | 51 (24.2) | 35 (14.7) | 14 (6.8) | 8 (3.8) |  |
| Some college | 208 (23.8) | 59 (28) | 56 (23.5) | 52 (25.2) | 40 (19) |  |
| College Degree | 223 (25.5) | 42 (19.9) | 65 (27.3) | 55 (26.7) | 60 (28.4) |  |
| Graduate School | 315 (36.1) | 50 (23.7) | 78 (32.8) | 85 (41.3) | 100 (47.4) |  |
| Other | 10 (1.1) | 5 (2.4) | 4 (1.7) | 0 (0) | 1 (0.5) |  |
| **Marital status** |  |  |  |  |  | 0.08 |
| Married | 443 (50.7) | 97 (46) | 121 (50.8) | 104 (50.5) | 118 (55.9) |  |
| No longer married | 372 (42.6) | 101 (47.9) | 108 (45.4) | 83 (40.3) | 76 (36) |  |
| Never married | 57 (6.5) | 13 (6.2) | 9 (3.8) | 18 (8.7) | 17 (8.1) |  |
| **Smoking status** |  |  |  |  |  | 0.26 |
| Current smoker | 25 (2.9) | 9 (4.3) | 9 (3.8) | 6 (2.9) | 1 (0.5) |  |
| Non-smoker | 492 (56.4) | 115 (54.5) | 140 (58.8) | 117 (56.8) | 118 (55.9) |  |
| Past smoker | 354 (40.5) | 86 (40.8) | 89 (37.4) | 83 (40.3) | 92 (43.6) |  |
| **Body mass index, kg/m<sup>2</sup>** | 27.6 ± 4.6 | 28.3 ± 4.6 | 27.6 ± 4.7 | 27.7 ± 4.4 | 26.8 ± 4.4 | **0.02** |
| **Multimorbidity index, 0 - 11** | 0.8 ± 0.9 | 1.1 ± 1.0 | 0.7 ± 0.8 | 0.8 ± 0.8 | 0.6 ± 0.7 | <.001 |
| **PFS Physical score, 0 - 50** | 15.8 ± 8.7 | 18.2 ± 9.5 | 16.2 ± 8.1 | 14.7 ± 8.3 | 14.1 ± 8.2 | <.001 |
| **PFS Mental score, 0 - 50** | 7.7 ± 7.8 | 10.5 ± 9.2 | 7.4 ± 7.0 | 6.7 ± 7.5 | 6.3 ± 6.3 | <.001 |
| **Total activity counts (100,000)\*\*** | 19.8 ± 5.9 | 18.0 ± 5.9 | 19.6 ± 6.0 | 20.7 ± 5.6 | 21.2 ± 5.4 | <.001 |
| **Average Daily Minutes of MVPA** | 186.4 ± 86.2 | 155.2 ± 85.8 | 183.4 ± 87.3 | 200.3 ± 80.6 | 211.6 ± 79.6 | <.001 |
| **MoCA, 13 - 30** | 24.8 ± 2.9 | 23.4 ± 3.2 | 24.9 ± 2.7 | 25.4 ± 2.5 | 25.8 ± 2.3 | <.001 |
| **CVLT Immediate Recall, 5 - 35** | 25.5 ± 4.4 | 23.8 ± 4.9 | 25.4 ± 4.3 | 26.0 ± 4.3 | 26.9 ± 3.7 | <.001 |
| **CVLT Delayed Recall, 0 - 9** | 6.0 ± 2.0 | 5.3 ± 2.2 | 6.0 ± 1.9 | 6.3 ± 2.0 | 6.5 ± 1.8 | <.001 |

Data are presented as mean ± SD for continuous variables and n (%) for categorical variables.
\**P*-value for continuous variables from the One-way ANOVA and Chi-Sq. goodness of fit test for categorical variables across severity strata.
The multimorbidity index includes self-report of physician diagnosis of: cancer (excluding nonmelanoma skin cancer), cardiac arrhythmia, chronic kidney disease, chronic obstructive pulmonary disease, coronary artery disease, congestive heart failure, dementia, depression, diabetes, stroke, and aortic stenosis.
\*\*Total activity counts represent the average 24-hour total activity imputed count, without the low frequency extension filter from the ActiGraph GT9X. PFS = Pittsburgh Fatigability Scale; MoCA = Montreal Cognitive Assessment; CVLT = California Verbal Learning Test; MVPA = moderate-to-vigorous physical activity

### Associations between Perceived Fatigability and Cognitive Function Assessments

After adjusting for demographics and multimorbidity index, we observed that for every 4-point higher PFS Physical score, the number of correct items on the DSST was lower by 0.7 items (β = -0.69, 95% confidence interval (CI): -1.09, -0.29). Associations between 3-point higher PFS Mental score and DSST were similar to PFS Physical (β = -0.64, 95% CI: -0.97, - 0.30) (Table 2). There were no statistically significant associations between perceived physical or perceived mental fatigability with MoCA (continuous or dichotomous) or CVLT Immediate or Delayed Recall (Table 2). Adjustment for total activity counts and mean daily minutes of MVPA slightly attenuated the associations, but they remained statistically significant (Table 2, models 3 and 4).

**Table 2.** Associations between Cognitive Function Assessments and Perceived Physical and Mental Fatigability: The Study of Muscle, Mobility and Aging (SOMMA) (N = 873)

|  | **4-point Higher PFS Physical** |  |  |  |
| --- | --- | --- | --- | --- |
|  | **Digit Symbol Substitution Test** | **Montreal Cognitive Assessment** | **CVLT Immediate Recall** | **CVLT Delayed Recall** |
| Model 1 | **-0.80 (-1.20, -0.41)** | 0.06 (-0.03, 0.14) | 0.00 (-0.14, 0.14) | 0.00 (-0.07, 0.06) |
| Model 2 | **-0.69 (-1.09, -0.29)** | 0.07 (-0.02, 0.16) | 0.01 (-0.13, 0.14) | 0.00 (-0.07, 0.06) |
| Model 3 | **-0.49 (-0.91, -0.07)** | 0.05 (-0.04, 0.15) | 0.05 (-0.10, 0.20) | 0.01 (-0.06, 0.08) |
| Model 4 | **-0.45 (-0.87, -0.03)** | 0.06 (-0.04, 0.15) | 0.05 (-0.09, 0.20) | 0.01 (-0.05, 0.08) |
|  | **3-point Higher PFS Mental** |  |  |  |
|  | **Digit Symbol Substitution Test** | **Montreal Cognitive Assessment** | **CVLT Immediate Recall** | **CVLT Delayed Recall** |
| Model 1 | **-0.76 (-1.09, -0.43)** | -0.02 (-0.09, 0.06) | -0.03 (-0.15, 0.08) | -0.02 (-0.08, 0.03) |
| Model 2 | **-0.64 (-0.97, -0.30)** | 0.00 (-0.08, 0.07) | -0.03 (-0.14, 0.09) | -0.02 (-0.08, 0.03) |
| Model 3 | **-0.52 (-0.87, -0.18)** | -0.01 (-0.08, 0.07) | 0.00 (-0.12, 0.12) | -0.01 (-0.07, 0.04) |
| Model 4 | **-0.50 (-0.84, -0.15)** | -0.01 (-0.08, 0.07) | 0.00 (-0.12, 0.13) | -0.01 (-0.07, 0.05) |

Results shown are beta coefficients and 95% confidence intervals.
Model 1: age + sex + race + education + marital status + site
Model 2: Model 1 + comorbidity index
Model 3: Model 2 + total activity counts
Model 4: Model 2 + MVPA
PFS = Pittsburgh Fatigability Scale; MVPA = moderate-to-vigorous physical activity
Total activity counts represent the average 24-hour total activity imputed count, without the low frequency extension filter from the ActiGraph GT9X.

### Effect Modification by Physical Activity

Physical activity volume and intensity modified the association of perceived mental fatigability with DSST score, both interactions *P* ≤ 0.01 (Table 3 and Figure 1). Though we only observed statistically significant associations in the lowest tertiles of average 24-hour total activity counts and average daily minutes of MVPA (i.e., the lowest volume of total activity and MVPA), there does appear to be a slightly graded, inverse relationship between lower volume and intensity of PA and stronger associations between perceived fatigability and cognitive function (both PFS subscales; Table 3 and Figure 1).

**Figure 1.**
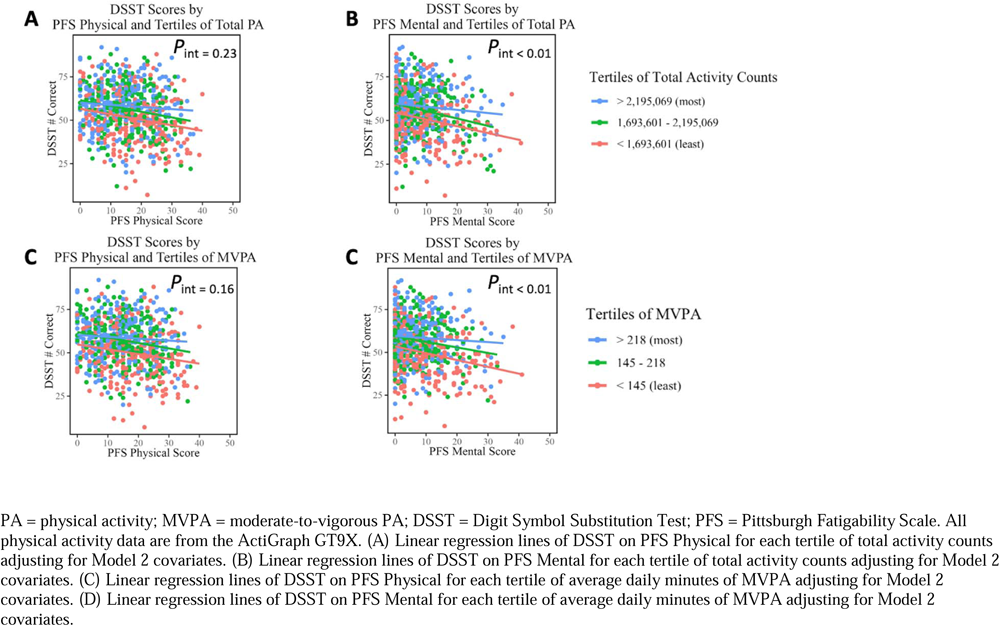
Effect Modification of the Association between Perceived Physical and Mental Fatigability and the Digit Symbol Substitution Test by Tertiles of Total Physical Activity (A & B) and Tertiles of Moderate-to-Vigorous Physical Activity (C & D): Study of Muscle, Mobility and Aging (SOMMA).

**Table 3.** Associations between the Digit Symbol Substitution Test and Perceived Physical and Mental Fatigability Stratified by Accelerometer-Measured Physical Activity: The Study of Muscle, Mobility and Aging (SOMMA)

|  | **PFS Physical score** | **PFS, Mental score** |
| --- | --- | --- |
| **Total Activity Counts,** |  |  |
| *P-interaction* | 0.23 | **<0.01** |
| Tertile 1 | **-0.87 (-1.59, -0.16)** | **-0.86 (-1.43, -0.28)** |
| Tertile 2 | -0.57 (-1.31, 0.18) | -0.59 (-1.22, 0.05) |
| Tertile 3 | -0.02 (-0.77, 0.73) | -0.05 (-0.66, 0.55) |
| **MVPA,** |  |  |
| *P-interaction* | 0.16 | **<0.01** |
| Tertile 1 | **-0.75 (-1.49, -0.01)** | **-0.86 (-1.43, -0.29)** |
| Tertile 2 | -0.55 (-1.25, 0.14) | -0.32 (-0.94, 0.29) |
| Tertile 3 | 0.04 (-0.73, 0.81) | 0.01 (-0.63, 0.64) |

PFS = Pittsburgh Fatigability Scale; MVPA = Average moderate-to-vigorous physical activity minutes per day; Counts = average 24-hour total activity vector magnitude counts
Results shown are beta coefficients and 95% confidence intervals.
All models are adjusted for age, sex, race, education, marital status, site, and comorbidity index.
MVPA Tertile 1: < 145 min/d; Tertile 2 ≥ 145 & < 218 min/d; Tertile 3 ≥ 218 min/d

## DISCUSSION

We provided evidence that greater perceived physical and mental fatigability were both significantly associated with worse performance on the DSST, but not with MoCA and either CVLT task. Additionally, we observed modification of the PFS Mental subscale – DSST association by both accelerometer-measured intensity and volume of PA metrics. Specifically, the associations of perceived mental fatigability with DSST were statistically significant and strongest in those with the lowest tertile of activity level (total activity counts and MVPA) after adjusting for demographics and the multimorbidity index.

There is limited literature to contextualize our findings. In a prospective cohort study of 2,781 community-dwelling older adults aged 65 – 94 without dementia that were enrolled in the Advanced Cognitive Training for Independent and Vital Elderly (ACTIVE) randomized intervention trial, Lin et al. found that participants who experience increased fatigue, measured by the Medical Outcomes Study 36-item Short-Form Health Survey (SF-36), over five years of follow up experienced significantly faster declines in memory, reasoning, and everyday speed composite scores relative to those with “persistent energy” (Lin et al., 2013). While Lin et al. used fatigue as the exposure, different cognitive function outcomes, and prospective latent class methods, the findings generally align with the present study: greater subjective fatigue and greater perceived physical and mental fatigability severity were both associated with poorer cognitive function. Additionally, the Long Life Family Study (N = 2,335; adults aged 60 – 108) found an association between higher DSST score (better performance) and lower PFS Physical score (less fatigability) (LaSorda et al., 2020). PFS Mental score was weakly associated with DSST score, but exhibited discrimination between higher vs. lower cognitive performance (LaSorda et al., 2020).

To further contextualize of our findings, we compared the strength of the fatigability– cognitive function association with the age–cognitive function association. After regressing DSST scores on Model 2 covariates omitting PFS score, the beta coefficient for 1-year higher age was -0.69 (β = -0.69, 95% CI: -0.87, -0.52), identical to that of the PFS Physical (β = -0.69, 95% CI: -1.09, -0.29) and nearly identical to PFS Mental (β = -0.64, 95% CI: -0.97, -0.30). It is well known that age is strongly associated with cognitive function and the present study provides evidence that the impact of fatigability is as strongly associated with cognitive function, independent of age.

Our stratified models contribute novel evidence that greater perceived physical and mental fatigability are more important at the lower end of the PA volume and intensity spectrum. Intervention studies on physical activity—mainly exercise—have shown that PA mitigates the deleterious effects of aging in the brain via the increasing of gray matter volume in frontal and hippocampal regions (Colcombe et al., 2006; Erickson et al., 2011; Mandolesi et al., 2018) and the modification of hippocampal biochemistry (Fernandes et al., 2017). In a study of the neural correlates of perceived fatigability in a sample of 29 adults enrolled in the Lifestyle Interventions and Independence for Elders Study (LIFE; mean age of 77.2 years), Wasson et al. provided initial evidence that those with higher perceived physical and mental fatigability had up to 3% less grey matter as a percentage of intracranial volume in certain regions of the brain compared to those with lower perceived physical and mental fatigability (Wasson et al., 2019). Those with more severe perceived fatigability may allow their perceptions to interfere with their activity, thus minimizing the potential neuroprotective effects of physical activity/exercise.

We hypothesized that we would observe associations between physical and mental fatigability with all the cognitive function assessments, but only identified associations with executive function/processing speed (as measured by the DSST) and not with memory. We believe this can be explained by two possible factors: 1) certain cognitive functions may have differential associations with fatigability, and 2) the healthier nature of the SOMMA participants masks the associations. First, there is a growing body of scientific evidence that indicates higher levels of physical activity/exercise are more strongly associated with executive function than other cognitive domains (Colcombe & Kramer, 2003; Hall et al., 2001), and these differential associations may extend to perceived fatigability given its association with physical activity/exercise. Understanding the relation between perceived fatigability and cognitive function may enrich our understanding of the PA and cognitive function relation as older adults may reduce their intensity or duration of activities to avoid feelings of fatigue (i.e., self-pace).

However, there do exist inconsistencies, as one meta-analytic review of randomized exercise intervention trials found no difference in the magnitude of the association between exercise with executive function but found a weaker association between exercise and short-term memory (Smith et al., 2010). Second, it is possible that the memory scores of the SOMMA participants are high enough to prevent the observation of any associations. Longitudinal studies with a more comprehensive neuropsychological battery are needed to conclude that there are no associations between perceived physical and mental fatigability with memory or general cognitive function. Lastly, examining whether performance fatigability, a measure of performance decrement during a task, is associated with these cognitive function assessments may elucidate the underlying mechanisms.

Our findings should be extended to a longitudinal framework to further understand the directionality of these associations. While SOMMA participants had no cognitive impairment at baseline, their levels of perceived fatigability may affect their performance on the cognitive function assessments and vice versa. As the participants included in the analytic sample were nearly exclusively White, generalizability of this study’s findings should be evaluated in more race/ethnicity diverse populations. The strengths of this study include the well-characterized cohort, large sample size of older adults, and high-quality measures of perceived fatigability, multiple cognitive function measures, and objectively assessed physical activity levels.

In conclusion, our study of over 850 older adults found that greater perceived physical and mental fatigability were associated with slower executive function and/or processing speed, but not memory. Individuals with greater perceived mental fatigability, especially those that also have the lowest levels of total activity and MVPA, might benefit from targeted interventions that reduce fatigability and in turn may beneficially influence cognitive function.

## Data Availability

All data produced in the present study are available upon reasonable request to the authors

https://sommaonline.ucsf.edu

## Acknowledgments

We would like to thank the SOMMA participants, research staff, and investigators. The analyses conducted in this manuscript were approved by SOMMA before conducting the analyses, but the analysis plan is not publicly available. SOMMA data is available upon request at https://sommaonline.ucsf.edu and the R code used to analyze the data for this manuscript is available upon request.

